# Integrative Mechanisms of Early Clinical and Research Training (ECART) in Orthopaedic Medical Education: A Qualitative Single-Case Study

**DOI:** 10.64898/2026.06.11.26355438

**Authors:** Yongfu Lou, Haoran Liu, Xiayue Xu, Yiming Xiao, Daoyi Ma, Wenyuan Shen, Chaoran Wang, Xiaohong Kong, Shiqing Feng

## Abstract

**Background:** Early clinical exposure and student participation in research are important components of medical training. They may support learning motivation, evidence literacy, and self-directed learning. In many programmes, however, clinical training and research training remain separated. Few studies have explained, within a real teaching team, how learners turn clinical phenomena into researchable questions and how research participation can reshape their clinical understanding. Early Clinical and Research Training (ECART) is a clinical-research integration approach developed by an orthopaedic team at the Second Hospital of Shandong University.

**Methods:** We conducted a theory-informed, interpretivist qualitative single-case study. The case was an orthopaedic clinical-research team at the Second Hospital of Shandong University. Participants included medical undergraduates, academic degree graduate students, professional degree graduate students, clinical teachers, and research platform leads. We used purposive sampling with maximum variation. Data were collected through semi-structured interviews and de-identified teaching documents. Data were analysed using the framework method and were interpreted with a Context-Activity-Mechanism-Outcome (CAMO) logic.

**Results:** The analysis showed that ECART was not simply early entry into the clinic or early entry into the laboratory. It was a team-based learning process centred on real medical problems. Four themes were identified. First, early clinical exposure helped learners make real problems visible and nameable, rather than merely increasing exposure. Second, clinical-research connection followed different pathways. Professional degree graduate students often started from clinical uncertainties in residency training and case management, and moved toward evidence-informed small projects. Academic degree graduate students often started from literature gaps, experimental findings, and mechanistic hypotheses, and then used clinical feedback to calibrate meaning. Third, research training, through literature reading, group meetings, experimental design, data review, and mentor questioning, helped learners move from completing tasks to explaining problems. Fourth, sustained ECART depended on a tiered team ecology formed by clinical teachers, research mentors, research platforms, and senior peers. Based on these findings, we refined the ECART programme theory: real medical problems are translated through explanation, searching, experimentalisation, and feedback-based reinterpretation into research questions that learners can understand, discuss, and test. This process supports problem formation, evidence awareness, mechanistic reasoning, translational judgement, and career clarification.

**Conclusion:** ECART is best understood as a clinical-research integrated learning ecology that emerges from real team practice, rather than as a fixed standardised course. Its educational value lies in a recurring cycle of real problems, research translation, multi-source feedback, and clinical reinterpretation. This framework may inform the design, evaluation, and contextual adaptation of clinical-research integration pathways in medical education.

## 1 Introduction

### 1.1 Trends in modern medical education

Modern medical education increasingly emphasises continuity across undergraduate education, graduate training, and continuing professional development. Medical learners need more than biomedical knowledge and clinical skills. They also need to develop problem awareness, evidence appraisal, research thinking, translational awareness, and a stable professional identity. As medical knowledge grows rapidly and clinical problems become more complex, classroom teaching and clinical skills training alone cannot meet the needs of advanced medical training. A key educational challenge is how to help learners understand problems, use evidence, conduct inquiry, and develop capacity for sustained growth in real medical contexts [1,2].

### 1.2 Separation of clinical and research training in the Chinese context

In China, medical education includes several training pathways. These include five-year undergraduate medical programmes, long-program or combined undergraduate-doctoral training, professional degree graduate training, and academic degree graduate training. Learners in these pathways have different goals. However, they often face a common problem: clinical training and research training are organised as separate domains. Clinical training tends to focus on patient care, diagnostic reasoning, clinical communication, and procedural skills.

Research training tends to focus on literature reading, laboratory participation, methodology, data analysis, and publication output. When these domains are not connected, learners may see clinical problems, literature evidence, research methods, and career development as separate tasks. They may struggle to build a continuous learning chain from seeing a clinical phenomenon, to raising a researchable question, and then to using research findings to reinterpret clinical practice [2].

This separation appears differently across pathways. For undergraduates, early clinical and research exposure may support interest and career exploration. Without guided questioning, however, it may remain at the level of observing scenes or trying techniques. For professional degree graduate students, clinical work, residency requirements, and examinations take up much of their time. Research training may become an extra burden if it is not connected to clinical uncertainty. For academic degree graduate students, laboratory work is the main activity. If their work lacks a clinical anchor, mechanistic research may drift away from real clinical needs.

Clinical-research integration should therefore not be reduced to earlier timing or adding more activities. It requires pathway-specific goals, tasks, and boundaries.

### 1.3 Gaps in existing research

Previous studies have shown that early clinical exposure can improve learning motivation, place basic medical knowledge in real contexts, and support professional identity formation [3-6]. Student participation in research can also develop critical thinking, information literacy, academic confidence, and self-directed learning [7-11]. Professional identity formation has also been highlighted as a key outcome in medical education research [12,13]. Yet most studies discuss “early clinical exposure” and “early research training” separately. Less is known about how these two forms of learning become connected in a real team-based learning environment.

Several questions need more detailed mechanistic explanation. How do learners identify researchable questions from clinical observation, case discussion, or diagnostic and therapeutic uncertainty? How do literature learning, group meetings, laboratory training, and mentor feedback work together to translate a clinical problem into a research hypothesis? How does research participation deepen learners’ understanding of clinical problems, evidence quality, disease mechanisms, and translational value? How do mentors, peers, and team ecology support learners from different pathways as they move between clinical and research domains? These gaps suggest the need to study clinical-research integrated learning in real team practice, with attention to context, activities, mechanisms, and boundary conditions [14-16].

### 1.4 Origin and meaning of ECART

In this study, Early Clinical and Research Training (ECART) was used as a practice-based concept to define the case. ECART originated from the long-standing educational practice of an orthopaedic clinical-research team at the Second Hospital of Shandong University. It refers to the connections among early clinical exposure, case discussion, literature learning, group meetings, research or laboratory training, mentor feedback, and peer support.

We did not treat ECART as a standardised, mature, or effectiveness-tested curricular intervention. Instead, we understood it as a practice-derived clinical-research integrated learning ecology. Within this ecology, learners engage with real cases, clinical questions, literature evidence, experimental design, data analysis, and feedback from multiple sources. They gradually learn how medical problems arise from clinical practice, how they can be translated into research questions, and how research findings can return to clinical interpretation and translational judgement. The core of ECART is not simply doing clinical work or research earlier. It is to make visible the learning process of clinical problems, research inquiry, and feedback-based reinterpretation.

### 1.5 Research questions

This study explored how early clinical exposure and research participation were connected within an orthopaedic clinical-research team. It focused on how learners from different training pathways integrated clinical observation and research inquiry in a real team environment. The study addressed four research questions:

#### Practice structure

What activities, learner roles, and organisational arrangements constitute ECART-related practice in an orthopaedic clinical-research team? How do these elements interact to support the integration of early clinical and research experiences?

#### Forward mechanism

How do learners from different training pathways move from clinical observation and clinical uncertainty to the identification and formation of research questions? What key steps and cognitive changes are involved?

#### Reverse mechanism

How does participation in research or laboratory platforms reshape learners’ understanding of clinical problems, evidence appraisal, mechanistic reasoning, and professional identity?

#### Boundary conditions

Which team contexts, mentor feedback patterns, peer support mechanisms, time pressures, and hierarchical relationships promote or constrain the implementation and sustainability of ECART?

Through these questions, we aimed to develop an initial programme theory explaining the contexts, activities, mechanisms, proximal outcomes, and boundary conditions of ECART. We also aimed to generate practical implications for the design and evaluation of clinical-research integration pathways.

## 2 Methods

### 2.1 Study design

We used an interpretivist, theory-informed qualitative single-case study design. The design was appropriate because the study aimed to explore how learners understood, experienced, and interpreted clinical-research integration in a real orthopaedic team. It was not intended to measure the effect size of a standardised intervention. A single-case study was suitable for analysing an educational process embedded in a specific team ecology and shaped by complex contextual factors. It also allowed us to develop an explanatory framework that may inform other contexts [17].

During analysis, ECART was treated as a sensitising concept derived from practice. We used the CAMO logic (Context-Activity-Mechanism-Outcome) as an interpretive heuristic. This helped us examine how specific activities activated learning mechanisms in particular contexts and led to proximal outcomes perceived by participants.

### 2.2 Study setting and case boundaries

The study setting was an orthopaedic clinical-research team at the Second Hospital of Shandong University. The team had access to orthopaedic clinical cases, research directions related to spine and spinal cord injury, research and laboratory platforms, mentor systems, and peer support. In this setting, ECART was defined as a set of connected learning practices. These included clinical observation, case discussion, literature learning, group meetings, research or laboratory training, research question refinement, and feedback from mentors and peers. The study focused on how these practices connected early clinical exposure with research participation.

The case boundary was defined by the setting, participant groups, educational activities, and mentoring relationships. Undergraduates engaged with ECART mainly through early clinical observation, case-based learning, and research introduction. Academic degree graduate students mainly used research tasks, literature appraisal, experiments, and methodological training to understand the research meaning of clinical problems. Professional degree graduate students developed research awareness from patient care, clinical uncertainty, and evidence use under the pressure of clinical work. Teachers and mentors provided explanations from the perspectives of clinical teaching, research platform management, project design, and team organisation.

### 2.3 Participants and sampling

Participants were medical education stakeholders who had participated in or guided ECART-related practice. They included medical undergraduates, academic degree graduate students, professional degree graduate students, and teachers or mentors. Inclusion criteria were: age 18 years or older; capacity to provide informed consent; participation in or guidance of ECART-related activities for at least six months, including clinical observation, case discussion, literature learning, research training, laboratory platform training, or mentor feedback; ability to discuss personal learning or teaching experience in an interview; and voluntary written informed consent. Exclusion criteria were: too little exposure to ECART; inability to complete the interview or refusal to be recorded; unwillingness to allow anonymised data to be used for analysis and publication; or a direct evaluative relationship with the recruiter that could not be adequately managed.

We used purposive sampling with maximum variation. We sought variation in training pathway, learning stage, duration of participation, team role, and viewpoint. We originally planned to interview 18 to 30 participants. The final sample size was determined by information power, theme repetition, the presence of negative cases, and the sufficiency of the analytical matrix. Recruitment stopped when additional interviews no longer changed the theme structure or CAMO interpretation in a meaningful way [18].

**Table 1.** Participant groups and main contributions.

| Participant group | n | Codes | Main experience sources | Analytical contribution |
| --- | --- | --- | --- | --- |
| Long-program medical undergraduates | 4 | U1-U4 | PBL, clerkship observation, outpatient or ward observation, laboratory introduction, group meetings, innovation projects | Early interest development, problem awareness, and career direction |
| Professional degree graduate students | 4 | P1-P4 | Clinical rotations, residency training, case management, database or small projects, evidence-based learning | Problem identification under clinical workload and lightweight research translation |
| Academic degree graduate students | 4 | A1-A4 | Laboratory training, literature reading, animal and cell experiments, mechanistic research, clinical handover meetings | How mechanistic research is calibrated by clinical value |
| Clinical teachers | 2 | C1-C2 | Ward rounds, case | How clinical |
|  |  |  | discussions, surgical teaching, clinical problem guidance | problems are raised, clarified, and transmitted |
| Laboratory PI / research platform mentors | 2 | PI1-PI2 | Platform management, project design, laboratory training, interdisciplinary feedback | How research platforms receive clinical problems and expose translational bottlenecks |

### 2.4 Data collection

Data were collected through semi-structured individual interviews. Each interview lasted approximately 20 to 30 minutes. The interview guide included common core questions and group-specific prompts for undergraduates, academic degree graduate students, professional degree graduate students, clinical teachers, and laboratory PI or research platform mentors. Interviews were conducted in quiet face-to-face settings when possible. Secure online interviews were used when needed. With participant consent, interviews were audio-recorded, transcribed verbatim, and de-identified.

Teaching documents were also collected to understand the case context and support data triangulation. These included anonymised training arrangements, learning task descriptions, meeting or group teaching materials, literature learning arrangements, and research training process documents. These documents were non-sensitive teaching materials. They were not used to measure student performance. We did not collect identifiable patient information, student grades, awards or penalties, assessment records, or other sensitive administrative data.

### 2.5 Researcher reflexivity

Some members of the research team had connections with the study setting. This helped the team understand local context, team practice, and professional language. It could also create risks of social desirability bias, over-familiarity, and selective interpretation. To reduce these risks, participants were told that participation was voluntary and would not affect grades, evaluation, resources, or teacher-student relationships. We avoided recruitment or interviewing by people with direct evaluative authority whenever possible. Interviewers wrote reflexive memos after interviews. During coding and interpretation, at least two researchers discussed highly interpretive data segments, coding disagreements, and negative cases.

### 2.6 Data analysis

Audio recordings were transcribed and de-identified before analysis. We used the framework method. First, two researchers read a subset of transcripts and documents independently. They recorded initial impressions and developed open codes. The research team then discussed these codes and developed an initial analytical framework. This framework included pre-specified dimensions such as early clinical observation, research participation, clinical-research connection, pathway differences, facilitators, barriers, boundary conditions, and perceived proximal outcomes. New themes were allowed to emerge inductively from the data [19].

Two researchers independently coded part of the dataset using this framework. They compared coding, discussed disagreements, and refined the framework. For data segments that were highly interpretive or open to different meanings, the team reached consensus through discussion. A third researcher was consulted when needed. After coding, we built a participant-category by theme by evidence matrix. We then integrated interview data, reflexive memos, negative case analysis, and document data to support interpretive synthesis.

In the interpretive stage, we used CAMO logic. Context referred to team ecology, training pathways, resources, and hierarchical relationships. Activity referred to clinical observation, case discussion, literature learning, research or laboratory platform training, mentor feedback, and peer support. Mechanism referred to potential mechanisms such as problem ownership, cognitive bridging, legitimate peripheral participation, identity expansion, and self-directed inquiry. Outcome referred to proximal educational outcomes such as problem formation, evidence awareness, mechanistic reasoning, clinical understanding, and self-positioning. Boundary conditions referred to constraints such as time pressure, role ambiguity, hierarchical silence, superficial clinical exposure, and publication pressure.

### 2.7 Trustworthiness and reporting standards

We enhanced trustworthiness through comparison across participant groups, triangulation between interviews and de-identified teaching documents, reflexive memos, dual coding discussions, negative case analysis, member reflection when appropriate, and a clear audit trail. Reporting was informed by SRQR and COREQ, especially regarding the research team, study design, sampling, interviews, analysis, reflexivity, and presentation of findings [20-22].

### 2.8 Ethics

The study was approved by the Research Ethics Committee of the Second Hospital of Shandong University (approval number: KYLL202605696). All participants provided written informed consent. Data were de-identified and handled according to privacy protection requirements.

## 3 Results

### Participant characteristics

Sixteen participants were included. They represented the main learner and mentor groups involved in ECART-related practice: four long-program medical undergraduates, four professional degree graduate students, four academic degree graduate students, two clinical teachers, and two laboratory PI or research platform mentors. All participants had participated in or guided activities related to clinical observation, case discussion, literature learning, research or laboratory training, group meetings, or mentor feedback.

Framework analysis generated four interrelated themes: early articulation of real clinical problems; pathway-specific clinical-research translation; the explanatory function of research participation; and tiered scaffolding with ecological governance.

#### Theme 1. Early articulation of real clinical problems and development of problem awareness

Data from undergraduates showed that the educational value of early clinical exposure was first reflected in making real medical problems visible. Through PBL, clerkship observation, outpatient observation, ward rounds, case discussion, and laboratory experience, students realised that diseases were not only defined by aetiology, pathology, and treatment principles. They also involved unresolved questions such as functional recovery, surgical improvement, inflammatory microenvironment regulation, AI-assisted diagnosis, and clinical translation. One undergraduate described a case of spinal cord injury as follows: “This case strengthened my confidence and determination to conduct research in this field…The case was a trigger, and the mentor was a bridge that connected us with research” (U1). Another student transformed the clinical problem into a more specific research focus: “The broad clinical question of why neurological function is difficult to restore after spinal cord injury was further translated into how acute inflammatory cells participate in secondary injury” (U3).

However, early exposure did not automatically lead to deep learning. Some students noted that without explanation and follow-up tasks, clinical exposure could remain at the level of seeing cases or feeling impressed. Laboratory training could also remain technical assistance. One undergraduate clearly distinguished meaningful training from formal participation: “ECART should not include activities without real problem awareness, such as simply visiting the laboratory, mechanically completing a single experimental step, or packaging concepts for competitions” (U3). Another student said that advanced content in large group meetings was often “highly scientific” for junior students, and without preparation “learning efficiency could be low” (U4). Thus, the key to early immersion was not earlier exposure itself. It was mentor explanation, literature tasks, and discussion feedback that helped students understand why what they saw constituted a medical problem.

#### Theme 2. Pathway-specific clinical-research translation mechanisms

Different training pathways entered ECART through different routes. Professional degree graduate students often started from concrete clinical uncertainties in residency training, rotations, and duty work. These included readmission after surgery, complications, prognostic differences, perioperative management, and evidence use. One professional degree graduate student noticed different causes of readmission after surgery for lumbar degenerative diseases during a spine surgery rotation. With mentor guidance, the student read previous work from the team and designed a small project: “This small project came directly from my observation and discovery in clinical practice” (P2). Another professional degree graduate student started from gastrointestinal dysfunction after lumbar surgery, reviewed literature on the gut-bone axis, and developed a project on lumbar degeneration and gut microbiota (P3). For professional degree graduate students, the key role of ECART was not to add another research burden. It was to translate uncertainty already present in clinical work into structured tasks that could be searched, discussed, designed, and fed back.

Academic degree graduate students followed a different route. They often started from literature gaps, experimental findings, mechanistic hypotheses, and mentor-led projects. They then used outpatient observation, clinical handover meetings, teacher explanations, and case feedback to calibrate the meaning of their research. One academic degree graduate student said: “I mainly work in the laboratory, and I have fewer opportunities for direct clinical contact than professional degree graduate students. I mainly learn about disease background and clinical pain points through mentors or clinical teachers in group meetings and project discussions” (A2). The same participant noted that ECART for academic degree graduate students was “not simply adding clinical observation, but establishing a connection mechanism among clinical problems, research training, and feedback-based correction” (A2). Therefore, academic degree graduate students did not need to be trained as professional degree clinicians. They needed sustained calibration of clinical value in early mechanistic research, so that laboratory work could be understood within real medical problems.

#### Theme 3. The explanatory function of research participation: from technical execution to mechanistic understanding

Research training became educationally meaningful when learners were asked to explain the origin of a problem, the value of the study, the meaning of indicators, and the pathway back to clinical practice. One undergraduate said that when preparing grant proposals, presentations, and defence materials, the student had to explain “what the clinical problem is, what the scientific hypothesis is, how the experiment verifies it, and what the results mean.” This process forced the student to be “not just doing experiments” (U3). One laboratory PI also stated that training outcomes should not be judged only by task completion, but by “thinking autonomy,” including whether students could connect experiments with clinical problems, explain experimental logic, and propose improvements based on results (PI1).

This explanatory training changed how learners understood clinical problems. Undergraduates moved from the view that it was enough to learn medical knowledge toward an understanding of the interdependence between clinical practice and research: “Before entering the team, I had no concept of research…Later, I found that research and clinical practice are inseparable” (U1). Professional degree graduate students moved from carrying out senior doctors’ plans toward active evidence searching and understanding of individual differences: “Previously I mainly followed treatment plans. After research training, I began to think about the reasons behind treatment plans and the advantages and disadvantages of different options” (P2). Academic degree graduate students moved from focusing on experimental indicators toward understanding disease complexity. One participant reflected: “After spending a long time in the laboratory, you may only focus on experimental indicators and paper figures, and sometimes lose sight of the original clinical problem” (A1). Another academic degree graduate student found that osteoporosis was not simply reduced osteogenesis. It involved osteogenic and adipogenic differentiation of bone marrow mesenchymal stem cells, osteoclasts, and the bone marrow microenvironment, which made the clinical problem “more complex and three-dimensional” (A4).

In contrast, when research training lacked clinical context, learners could experience it as technical labour, publication work, or a split between learning and use. One professional degree graduate student noted that some basic research focused only on rare molecules or signalling pathways and did not consider clinical accessibility or translational potential. Such work could gradually become detached from clinical scenes (P1). This suggests that the educational focus of ECART is not to increase the number of research activities. It is to ensure that each research activity responds to a clear clinical problem.

#### Theme 4. Tiered scaffolding and ecological governance: support conditions and boundaries for sustainable ECART

ECART depended on a scaffold formed by clinical teachers, research mentors, laboratory PI or research platform mentors, and senior peers. Clinical teachers raised, clarified, and explained real clinical problems. One clinical teacher noted that professional degree graduate students were willing to solve diagnostic and therapeutic problems, but their weakness was that they “did not know how to refine these problems, summarise these problems, or explore them through research” (C1). Research mentors translated problems into hypotheses, technical routes, and evidence chains, and also safeguarded research integrity. Another clinical teacher emphasised that research mentors should not only guide methods but also review all experimental data and continually emphasise research integrity (C2). Research platforms functioned as translators of clinical problems and exposers of translational bottlenecks. PI2 described the platform as “a translator of clinical problems, an exposer of translational bottlenecks, and a convener of interdisciplinary feedback.” Senior peers lowered the technical and psychological entry threshold through experimental demonstration, literature experience, time-management advice, and emotional support.

This team ecology had to operate in a tiered way. Undergraduates needed interest development, problem awareness, and experience of the full research process. Professional degree graduate students needed lightweight, problem-oriented, evidence-based clinical research with clear ethical boundaries. Academic degree graduate students needed clinically anchored mechanistic innovation rather than simply more clinical tasks. PI1 explicitly stated that the core goal for undergraduates was initiation and cognition, academic degree graduate students should focus on mechanistic innovation, and professional degree graduate students should emphasise application and translation. At the same time, multi-source feedback carried risks. PI1 noted that senior students might have experience-based bias, which could lead to errors in knowledge transmission. PI2 noted that a PI’s own views might become outdated, while students might not dare to question or challenge them. ECART therefore requires standardised group meetings, case discussions, staged reflection, and open discussion spaces to reduce fragmented feedback and dependence on authority.

### Revised ECART programme theory

Based on these themes, we refined the ECART programme theory. In a team context with real clinical cases, research platforms, mentorship, and peer scaffolding, clinical problems are translated through explanation, searching, experimentalisation, and feedback-based reinterpretation into research questions that learners can understand, discuss, and test. This process activates problem ownership, cognitive bridging, legitimate peripheral participation, and clinical-research identity expansion. It then supports proximal outcomes such as problem formation, evidence awareness, mechanistic reasoning, translational judgement, and career clarification. Effective ECART does not depend on a single curricular template. It depends on tiered goals, continuous feedback, open communication, and clinical value calibration.

**Figure 1.**
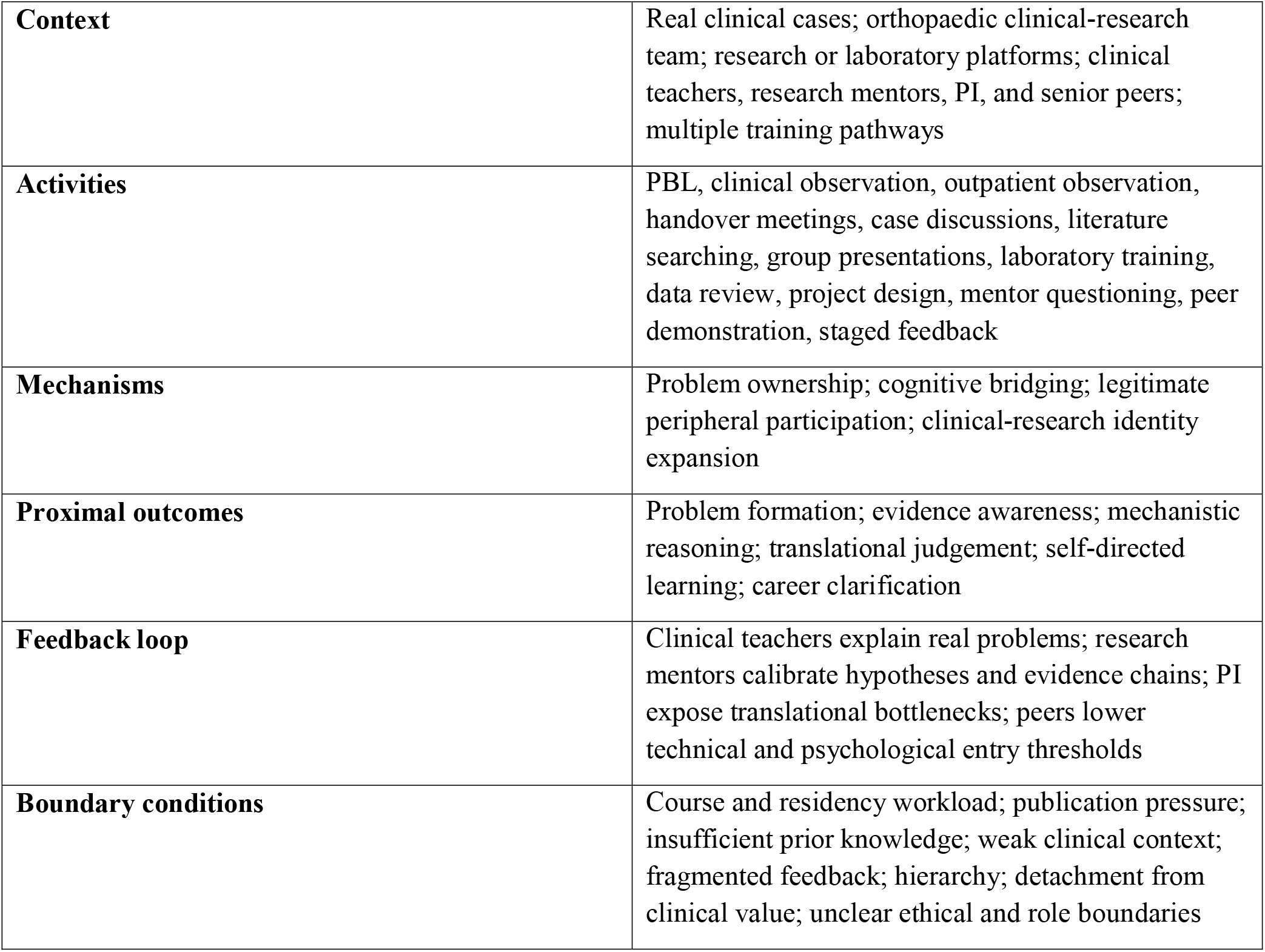
Logic of the revised ECART programme theory

## 4 Discussion

### 4.1 Main findings and contributions

This study developed an explanatory understanding of how ECART operated in an orthopaedic clinical-research team. The findings show that the connection between early clinical exposure and research participation did not occur automatically. It was also not dependent on a single course design. Instead, it was gradually formed through sustained interactions among real cases, literature evidence, laboratory platforms, mentor feedback, and peer scaffolding. Through these interactions, learners moved from seeing clinical phenomena, to forming researchable questions, and then to using research to reinterpret clinical practice.

The first finding is that real clinical problems must be educationally translated before they become learning resources. Orthopaedic conditions, especially spinal cord injury, lumbar degeneration, postoperative complications, and functional recovery problems, are complex, long-term, and strongly linked to translational needs. These problems can stimulate interest. However, only through mentor explanation, literature searching, case review, and questioning in group meetings can they become research problems that can be discussed and tested. The second finding is that learners from different pathways do not share one single clinical-research connection. Undergraduates mainly experienced interest development and career clarification. Professional degree graduate students mainly translated clinical uncertainty into evidence-informed small projects. Academic degree graduate students mainly recalibrated mechanistic research toward clinical value. The third finding is that the educational value of research training lies not only in technical learning or publication output. It also lies in training learners to explain problem origins, evidence chains, mechanistic hypotheses, and translational boundaries. The fourth finding is that ECART depends on a tiered team ecology. Clinical teachers provide real problems. Research mentors translate methods. Research platforms expose translational bottlenecks. Senior peers lower technical and psychological entry barriers.

This study contributes to the literature in three ways. First, it does not discuss early clinical exposure and early research training as separate educational activities. It explains how they are connected, translated, and fed back in a real team ecology. Second, it reveals differences in ECART mechanisms across training pathways. This avoids treating undergraduates, professional degree graduate students, and academic degree graduate students as if they required the same training model. Third, from an orthopaedic setting, the study proposes an initial programme theory of real problems, research translation, multi-source feedback, and clinical reinterpretation. This theory can be tested and adapted in future studies of clinical-research integrated education.

### 4.2 Relationship with previous research

Previous studies on early clinical exposure have shown that real clinical contexts can enhance motivation, contextualise knowledge, and support professional identity formation. Our study is consistent with these findings, but it adds that the educational effect of early exposure depends less on the timing of exposure itself and more on whether the clinical phenomenon is interpreted as a problem. In this study, observation, visits, or attendance alone were not enough for deep learning. Early exposure was more likely to become problem awareness and inquiry motivation when case phenomena, clinical uncertainty, and knowledge gaps were made explicit and connected with literature and research tasks [3-6,12,13].

Research on medical student research participation has often emphasised critical thinking, information literacy, academic confidence, and publication-related skills. Our study further highlights the clinical return pathway of research training. Through literature reading, experimental design, model selection, and data review, learners not only acquired research methods. They also reinterpreted the complexity of clinical problems, evidence quality, and mechanistic boundaries. This suggests that research training should not be assessed only by technical competence, project completion, or publications. It should also be assessed by whether learners can explain where the research question came from, what clinical pain point the indicators respond to, and whether the findings can explain or improve a real clinical problem [7-11].

This study also adds a China-specific explanation of clinical-research integration under multiple training pathways. Unlike some curricular or standardised integration models, ECART in this study was closer to a tiered learning ecology in team practice. Its value lies not in designing the same task for all learners. Instead, it lies in designing different forms of clinical input, research tasks, and feedback according to learners’ time structure, prior knowledge, and training goals [2].

### 4.3 Implications for educational practice

The first practical implication concerns the selection and organisation of real problems in orthopaedics. Orthopaedic conditions often involve imaging interpretation, surgical decision-making, postoperative rehabilitation, functional assessment, complication management, and mechanisms of tissue repair. These issues are suitable triggers for clinical-research integrated learning. Teachers can build a case-literature-mechanism-experiment-feedback chain around problems such as functional recovery after spinal cord injury, readmission after surgery for lumbar degeneration, perioperative complications, osteoporosis mechanisms, scar formation, and tissue repair. Such a chain can help learners understand how problems arise from clinical practice and how research methods can break them down.

For undergraduates, ECART should focus on interest development, problem awareness, and experience of the research process. It should not require students to take on complex independent projects too early. Useful tasks include literature searching after PBL, problem logs after outpatient observation or case discussion, preparatory reading before group meetings, explanations that link basic laboratory steps to research aims, and small research case design exercises. It is especially important to avoid limiting undergraduates to mechanical assistance or competition packaging. Staged reflection should help them understand where each task fits in the full research chain.

For professional degree graduate students, ECART should support clinical competence rather than compete with residency training. Suitable forms include case-driven literature presentations, follow-up or readmission database analysis, perioperative management review, small-sample clinical question studies, guideline and evidence discussions, and short-cycle quality improvement projects. The key is not to increase basic laboratory burden. It is to help professional degree graduate students translate recurring clinical uncertainties into feasible, evidence-based, and ethically clear inquiry tasks.

For academic degree graduate students, ECART should not mean adding many clinical duties. It should mean building a stable clinical anchoring mechanism. This can include regular attendance at orthopaedic handover meetings, clinical-research joint group meetings, outpatient observation with mentors, clinical teachers’ explanations of disease background, and case feedback on research hypotheses. Research training for academic degree students should ask: why is this mechanism being studied, can this model represent the clinical problem, and how can the experimental result return to disease understanding and translational judgement? These questions can reduce the risk that basic research becomes detached from real clinical needs.

For team leaders, ECART requires institutionalised scaffolding rather than reliance on individual enthusiasm. Clinical teachers, research mentors, PI, and senior peers should have clear roles. Clinical teachers provide and explain real problems. Research mentors translate methods and review evidence chains. PI or platform mentors integrate resources and identify translational bottlenecks. Senior peers provide daily technical guidance and experience transfer. Standardised group meetings, unified technical training, research integrity education, staged reporting, and open discussion can reduce fragmented feedback, experience-transfer bias, and hierarchical silence [14-16].

### 4.4 Strengths and limitations

A strength of this study is that it used a real orthopaedic clinical-research team as the case. It included multiple perspectives from undergraduates, professional degree graduate students, academic degree graduate students, clinical teachers, and laboratory PI or research platform mentors. The framework method and CAMO logic allowed us to move beyond describing activities. We explained how these activities activated or constrained learning mechanisms in different pathways. This led to an initial programme theory with mechanistic explanatory value.

This study also has limitations. First, the sample size was 16. Although it covered five key participant groups, each group was small. In particular, there were only two clinical teachers and two PI or research platform mentors. This may not fully capture different teaching styles, platform management models, or negative cases. Second, the study was conducted in a single centre and one team. The case focused on orthopaedics, especially spine and spinal cord injury research. Orthopaedics has visually accessible cases, rich imaging and surgical scenes, and strong needs for functional recovery and translation. The mechanisms identified here may be most transferable to surgical or translational medicine settings that have clinical case resources, research platforms, and mentoring teams. They may not apply directly to disciplines with fewer resources, different case types, or limited research platforms. Third, the data came mainly from interviews and team documents. They revealed participants’ experiences and meaning-making, but they also relied on recall and self-explanation. Social desirability bias may remain. Fourth, the study focused on proximal educational outcomes, such as problem awareness, evidence awareness, mechanistic reasoning, and career clarification. It did not track whether these changes led to long-term research competence, clinical reasoning, or career development.

The transferability of this study should therefore be understood at the level of mechanisms, not as a direct curriculum package. For medical education settings with stable mentorship, clinical case resources, research platforms, and peer scaffolding, the mechanism of real problems, research translation, multi-source feedback, and clinical reinterpretation may be useful. In settings with weak feedback, superficial clinical exposure, insufficient research resources, or unsafe hierarchical communication, ECART may operate differently and produce different outcomes.

### 4.5 Future research

Future research can proceed in three directions. First, multi-case studies across departments and hospitals can examine which elements of ECART are stable across contexts and which depend on orthopaedics, spinal cord injury research, or specific team resources. Second, longitudinal studies can follow learners from undergraduate to graduate stages, or from early team entry to independent project work, to observe changes in problem awareness, evidence use, mechanistic reasoning, and professional identity. Third, mixed-methods studies can develop and evaluate process and proximal outcome indicators for ECART. These may include the quality of clinical question generation, use of literature evidence, clarity of research hypotheses, quality of group feedback, ability to reinterpret clinical problems, and translational judgement.

## 5 Conclusion

Based on a qualitative single-case study of an orthopaedic clinical-research team, this study reached three core conclusions. First, the value of early clinical exposure and research participation does not lie in earlier exposure alone. It lies in a recurring cycle of real problems, research translation, multi-source feedback, and clinical reinterpretation. This cycle helps learners translate clinical phenomena into research questions that can be understood, discussed, and tested.

Second, ECART operates differently across training pathways. Undergraduates mainly need interest development and experience of the research process. Professional degree graduate students need to translate clinical uncertainty into evidence-informed small projects. Academic degree graduate students need to calibrate mechanistic research against clinical value.

Third, ECART depends on a team ecology. Clinical teachers, research mentors, platform leads, and senior peers provide problem clarification, methodological translation, resources, and psychological and technical support. These supports are essential for sustainable clinical-research integrated learning.

Grounded in China’s multi-track medical training context, this study provides a locally relevant mechanism and practical ideas for building clinical-research integrated teaching systems in orthopaedics and similar clinical specialties. It highlights the need for pathway-specific tasks, embedded clinical context, and continuous feedback across training stages.

## Data Availability

The de-identified raw interview transcripts are not publicly available to protect participant privacy. Selected de-identified analysis materials may be made available from the corresponding author upon reasonable request and subject to institutional ethics approval.

## Declarations

### Ethics approval and consent to participate

This study was approved by the Research Ethics Committee of the Second Qilu Hospital of Shandong University (approval number: KYLL202605696). All participants were informed of the study purpose, procedures, and data use. Written informed consent was obtained from all participants. Interview data and personal information were de-identified throughout the study. Participants could stop the interview or decline to answer any question at any time.

### Consent for publication

Not applicable. This manuscript does not contain identifiable personal information.

### Availability of data and materials

To protect participant privacy, the de-identified raw interview transcripts are not publicly available. Selected analysis materials may be made available after institutional ethics review and approval by the corresponding author.

### Authors’ contributions

Study conception and design and interview guide development(Shiqing Feng, Xiaohong Kong). Interview implementation, audio collection, and field coordination(Yongfu Lou, Haoran Liu). Transcription, transcript cleaning, terminology checking, and formatting(Xianyue Xu, Yiming Xiao, Daoyi Ma). Data coding, analysis, result synthesis(Yongfu Lou, Haoran Liu), manuscript drafting, and revision(Wenyuan Shen, Chaoran Wang). All authors discussed the study, reviewed the final manuscript, and approved it for submission.

## Acknowledgements

Not applicable.

## References

[1] Frenk J, Chen L, Bhutta ZA, et al. Health professionals for a new century: transforming education to strengthen health systems in an interdependent world. Lancet. 2010;376(9756):1923–1958.

[2] Liu X, Jiang J, Li Y, et al. Medical education systems in China: development, status, and evaluation. Academic Medicine. 2023;98(1):43–51.

[3] Dornan T, Littlewood S, Margolis SA, et al. How can experience in clinical and community settings contribute to early medical education? A BEME systematic review. Medical Teacher. 2006;28(1):3–18.

[4] Yardley S, Teunissen PW, Dornan T. Experiential learning: AMEE Guide No. 63. Medical Teacher. 2012;34(2):e102–e115.

[5] Peters S, Clarebout G, Diemers A, et al. Enhancing the connection between the classroom and the clinical workplace: a systematic review. Perspectives on Medical Education. 2017;6(3):148–157.

[6] AkbariRad M, Khadem-Rezaiyan M, Ravanshad S, et al. Early clinical exposure as a highly interesting educational program for undergraduate medical students: an interventional study. BMC Medical Education. 2023;23:292.

[7] Lee GSJ, Chin YH, Jiang AA, et al. Teaching medical research to medical students: a systematic review. Medical Science Educator. 2021;31(2):945–962.

[8] Murray H, Savage T, Hilton J, et al. Research training during medical school: a scoping review. Medical Science Educator. 2022;32(6):1583–1595.

[9] Carberry C, McCombe G, Tobin H, et al. Curriculum initiatives to enhance research skills acquisition by medical students: a scoping review. BMC Medical Education. 2021;21:312.

[10] Riiser K, Kalleson R, Holmen H, et al. Integrating research in health professions education: a scoping review. BMC Medical Education. 2023;23:653.

[11] Stone C, Dogbey GY, Klenzak S, et al. Contemporary global perspectives of medical students on research during undergraduate medical education: a systematic literature review. Medical Education Online. 2018;23(1):1537430.

[12] Cruess RL, Cruess SR, Boudreau JD, et al. Reframing medical education to support professional identity formation. Academic Medicine. 2014;89(11):1446–1451.

[13] Findyartini A, Greviana N, Felaza E, et al. Professional identity formation of medical students: a mixed-methods study in a hierarchical and collectivist culture. BMC Medical Education. 2022;22:443.

[14] Ramani S, Kusurkar RA, Lyon-Maris J, et al. Mentorship in health professions education: an AMEE Guide for mentors and mentees (AMEE Guide No. 167). Medical Teacher. 2024;46(8):999–1011.

[15] Toh RQE, Koh KK, Lua JK, et al. The role of mentoring, supervision, coaching, teaching and instruction on professional identity formation: a systematic scoping review. BMC Medical Education. 2022;22:531.

[16] King E, Turpin M, Green W, et al. Learning to interact and interacting to learn: a substantive theory of clinical workplace learning for diverse cohorts. Advances in Health Sciences Education. 2019;24(4):691–706.

[17] Yin RK. Case study research and applications: design and methods. 6th ed. Thousand Oaks: Sage; 2018.

[18] Malterud K, Siersma VD, Guassora AD. Sample size in qualitative interview studies: guided by information power. Qualitative Health Research. 2016;26(13):1753–1760.

[19] Gale NK, Heath G, Cameron E, et al. Using the framework method for the analysis of qualitative data in multidisciplinary health research. BMC Medical Research Methodology. 2013;13:117.

[20] O’Brien BC, Harris IB, Beckman TJ, et al. Standards for reporting qualitative research: a synthesis of recommendations. Academic Medicine. 2014;89(9):1245–1251.

[21] Tong A, Sainsbury P, Craig J. Consolidated criteria for reporting qualitative research (COREQ): a 32-item checklist for interviews and focus groups. International Journal for Quality in Health Care. 2007;19(6):349–357.

[22] Stenfors T, Kajamaa A, Bennett D. How to assess the quality of qualitative research. Clinical Teacher. 2020;17(6):596–599.

